# Transcriptomic and connectomic correlates of differential spatial patterning among glioblastomas and low-grade gliomas

**DOI:** 10.1101/2022.07.04.22277220

**Authors:** Rafael Romero-Garcia, Ayan S. Mandal, Richard AI Bethlehem, Benedicto Crespo-Facorro, Michael G Hart, John Suckling

## Abstract

**Background:** Unravelling the complex events driving grade-specific spatial distribution of brain tumour occurrence requires rich datasets from both healthy individuals and patients. Here, we combined open-access data from The Cancer Genome Atlas, the UKBiobank and the Allen Brain Human Atlas to disentangle how the different spatial occurrences of Glioblastoma Multiforme (GBM) and Low-Grade Gliomas (LGG) are linked to brain network features and the normative transcriptional profiles of brain regions.

**Methods:** From MRI of brain tumour patients we first constructed a grade-related frequency map of the regional occurrence of LGG and the more aggressive GBM. Using associated mRNA transcription data, we derived a set of differential gene expressions from GBM and LGG tissues of the same patients. By combining the resulting values with normative gene expressions from *postmortem* brain tissue, we constructed a grade-related expression map indicating which brain regions express genes dysregulated in aggressive gliomas. Additionally, we derived an expression map of genes previously associated with tumour subtypes in a GWAS study (tumour-related genes).

**Results:** There were significant associations between grade-related frequency, grade-related expression, and tumour-related expression maps, as well as functional brain network features (specifically, nodal strength and participation coefficient) that are implicated in neurological and psychiatric disorders.

**Conclusions:** These findings identify brain network dynamics and transcriptomic signatures as key factors in regional vulnerability for GBM and LGG occurrence, placing primary brain tumours within a well-established framework of neurological and psychiatric cortical alterations.

## INTRODUCTION

Adult diffuse gliomas are devastating and lethal types of cancer. According to the World Health Organization (WHO) grading system, survival rates drastically vary from Low-Grade Gliomas (LGG) with median survival between 4.7 and 9.8 years^1^ to Glioblastoma Multiforme (GBM), or grade IV astrocytoma, with survival limited to around 15 months.^2^ Brain tumours occur via several aetiologies and are located heterogeneously across the brain. Although the location of the tumour determines the likelihood of complete resection, and thus long-term survival^3^, its specific location can influence the accompanying cognitive changes that patients often experience. In this study, we leverage well-established open access data sources to construct a map of the distribution of brain tumours of varying grade, and investigate the reasons for these distributions in the underlying genetic expression and functional networks. LGGs, most frequently astrocytomas and oligodendrogliomas, are slow-growing, infiltrative tumours that account for 10-20% of all primary brain tumours.^1^ Most, usually younger patients, will die due to the malignant (anaplastic) transformation of the tumour to a higher grade. The rate of malignant transformation is diverse amongst patients, and usually determined radiologically. GBM is the most common type of primary malignant brain tumour. In about ∼90% of cases, they develop *de novo* as primary tumours with a high grade without histologic evidence of a precursor lesion.^4^

The traditional grading system based on histological appearance does not always reflect the biological behaviour of tumours, and in 2016 WHO incorporated a molecular classification criterion for adult diffuse gliomas.^5^ Recent investigations have revealed that tumour molecular profiles have a greater prognostic value and are better predictors of tumour growth kinetics, regardless of grade or histology.^6,7^ For example, mutations of Telomerase Reverse Transcriptase (TERT) and Isocitrate Dehydrogenase 1 (IDH1-M), methylation of the O-6-methylguanine-DNA methyltransferase gene (MGMT-met) and codeletion of chromosomes 1p and 19q (1p19q-cod) lead to germline variants that are driven by distinct pathogenic mechanisms characterised by specific proliferation rates and aggressiveness. Molecular signature also influences tumour location, with IDH-M gliomas preferentially located in the frontal lobe adjacent to the rostral extension of the lateral ventricles.^8^ It has been hypothesised that the high glutamate flux present in prefrontal cortex creates a metabolic niche that supports IDH-M gliomas.^9^ Beyond these well-known genetic mutations, a GWAS including >12,000 glioma patients and 18,000 controls identified 25 risk loci associated with glioma risks in adults.^10^ These tumour-related genes are heterogeneously distributed across glioma subtypes and grades, suggesting that they also play a major role in tumour appearance and progression.^11^

At the end of the Nineteenth Century, Paget described the seed and solid hypothesis that successful tumour growth depends on interactions between the properties of cancer cells (seeds) and their potential target tissue (soil).^12,13^ Although this hypothesis has been extensively explored as a conceptual scaffold for understanding tumour metastasis, we propose its re-examination in the context of the cellular origins of primary gliomas and their progression.

It has been hypothesised that gliomas originate from neurogenic niches of Neuronal-Stem Cells (NSCs) and Oligodendroctye Precursor Cells (OPCs) in the subventricular zone^14^ that migrate along large-scale axonal tracts to populate distributed cortical areas.^15^ Neural-glioma cellular interactions are key determinants of glioma growth and migration^16^ forming a positive feedback loop by which glioma progression is promoted by molecules secreted in neuronal communication^17^, and increased glutamate release from gliomas inducing hyperexcitability of cortical networks.^18^ We recently demonstrated that regions with a high number of functional connections (i.e., hubs) and with elevated participation coefficient (i.e., regions interconnecting constituent network communities) are more vulnerable to the instantiation of gliomas.^19^ Collectively, this evidence suggests that structural and metabolic factors are key determinants in tumour progression. However, it neglects the potential contribution of molecular and transcriptomic factors. Here, we investigate whether the cellular and gene expression profiles of the brain regions where glioma cells migrate are related to grade-specific occurrence.

International collaborative efforts have not just generated genomic, epigenomic, transcriptomic, proteomic and neuroimaging data, but have also provided publicly accessible platforms to a growing research community to advance our understanding of the molecular basis of glioma. Datasets incorporating molecular and radiological information are particularly valuable for connecting genotypic and phenotypic profiles. A representative example of this effort is The Cancer Genome Atlas (TCGA)^20^ which, in coordination with the Cancer Imaging Archive (TCIA)^21^, recruited LGG and GBM patients and gathered transcriptomic data from tumour tissue and MRI scans from the same patients. Despite the unquestionable utility of this resource, gene expression in tumour tissue largely diverges from that in normal tissue, which can be collected from near the tumour, but only in a limited number of cases. Complementarily, the Allen Human Brain Atlas (AHBA) currently has the most exhaustive spatial coverage of gene expression data in brain tissue derived from six *post-mortem* normative donors.^22^ Despite these donors having had no brain disease, their spatially resolved gene expression profiles are useful in shedding light on the transcriptomic vulnerabilities of the different brain areas.^23–25^

In this study, we exploited the TCGA dataset to identify genes differentially expressed in GBM and LGG, and combine these expressions with the ABHA to construct a map of grade-related expression across the entire cortex from normative control data. We additionally incorporated functional connectivity data from the UK Biobank (UKB), one of the most ambitious MRI studies to date.^26^ We hypothesized that the spatial profile of grade-related occurrence would be associated with grade-related expression topography. Moreover, we also expected this profile to be associated with tumour-related gene expression and functional brain network attributes, revealing a grade-sensitive pattern of regional vulnerability to brain tumours.

## METHODS

### TCGA MRI brain tumour masks

The TCGA dataset (https://www.cancer.gov/about-nci/organization/ccg/research/structural-genomics/tcga/studied-cancers/glioblastoma) includes solid samples of GBMs and LGGs of which 135 and 107, respectively, were matched to MRI scans from The Cancer Imaging Archive (TCIA) dataset (https://wiki.cancerimagingarchive.net/display/Public/Brain-Tumor-Progression) collected at 13 institutions. Diagnoses of GBM and LGG were established at the contributing institutions and reviewed by neuropathologists in the TCGA consortium^20,27^.

T1-weighted MRIs were skull-stripped, co-registered and resampled to 1 mm^3^ resolution. The segmentation algorithm GLISTRboost was used to classify voxels into four categories: contrast-enhancing tumour, necrotic non-enhancing core, peritumoural oedema and normal brain tissue. Labels were then manually corrected by board-certified neuroradiologists.^28^ Group-level GBM and LGG occurrence was obtained by concatenating glioma masks across patients. Frequency maps were first calculated at the voxel level as the proportion of times that a given voxel was overlapped by a tumour mask, and then at regional level as the average overlapping of all voxels within each parcel of an atlas. See yellow box in **Figure 1** for a summary.

**Figure 1:**
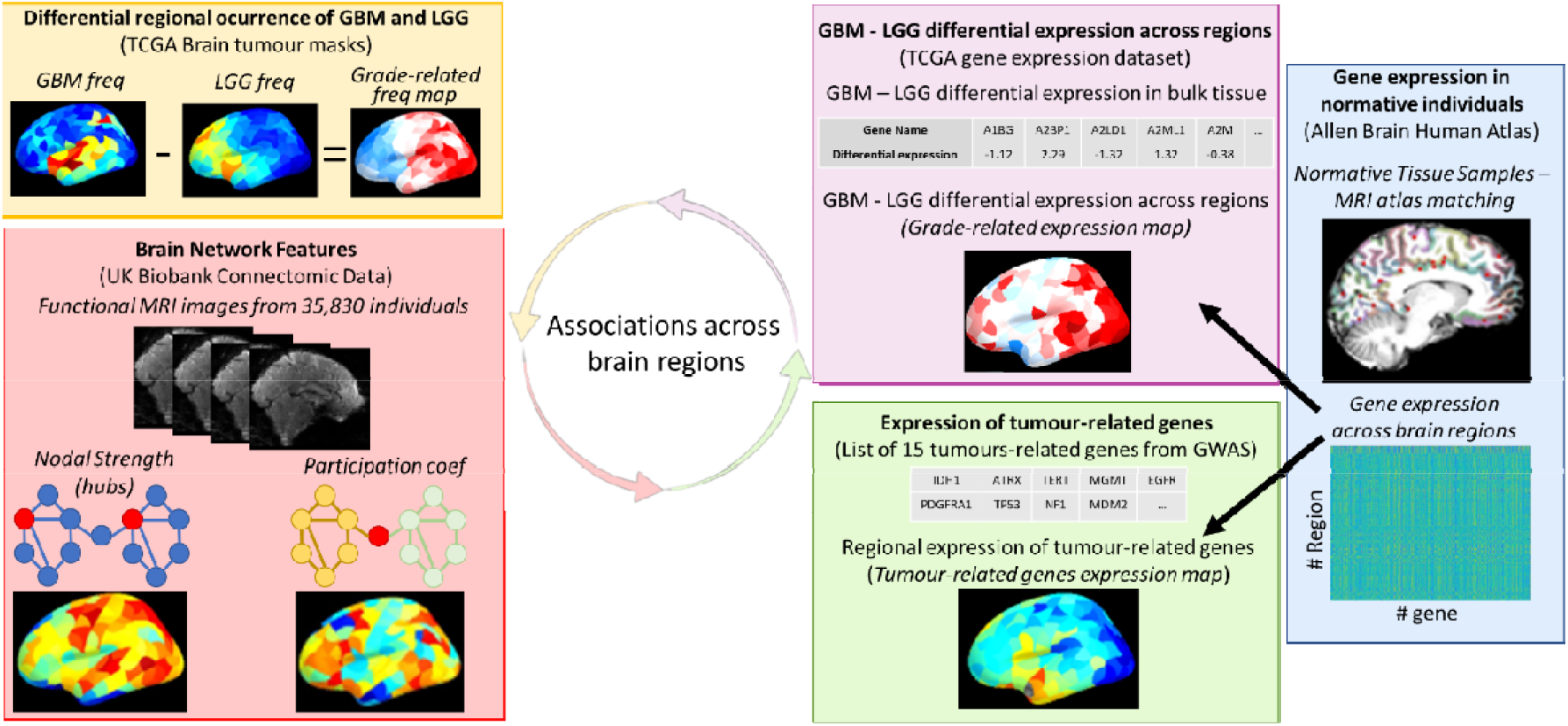
Flowchart of data processing and analysis. Grade-related frequency maps were derived from the TCGA brain tumour masks (yellow box) and brain network features were extracted from the UK Biobank fMRI dataset (red box). Grade-related expression regional values (purple box) were calculated by combining the differential expression in bulk tumour tissue with the ABHA (blue box). The spatial expression pattern of 15 genes from a previous GWAS glioma study^11^ was computed from the ABHA (green box).

### TCGA bulk transcriptomic analysis

Following a previously reported workflow,^29^ we derived bulk mRNA transcripts of the whole genome (14,899 genes after exclusions) from the N=672 individuals from the TCGA dataset. Using the edgeR package,^30^ differential expression analyses compared tissue from patients with GBM (n=156) and LGG (n=516). As a result, we obtained a log count per million differentiation index per gene where positive values indicate higher expression in GBM compared to LGG bulk tissue. This pipeline was additionally performed by comparing IDH wild-type (n=124) and IDH mutated (n=88) bulk tissue to derive differential expression values to establish potential similarities with the grade-related expression. This pre-processing is described in more detail elsewhere.^31^ Enrichment for cellular and biological components of the resulting differentially expressed genes were assessed using Enrich (https://maayanlab.cloud/Enrichr/).^32^

### ABHA normative gene expression map

The ABHA provides microarray expression data from six normative donors sampled from over 3,000 locations that covered the whole left hemisphere and were analysed using >62,000 probes per profile.^22^ As the spatial location of the samples was annotated and brains were scanned prior to tissue resection, it is possible to match a gene expression profile with a specific brain area.^33^ The dataset was preprocessed using standard protocols implemented in the Abagen toolbox (https://abagen.readthedocs.io/)^34^: (*i*) microarray probes were reannotated (probes not matching a valid Entrez ID were discarded), (*ii*) probes with expression intensity less than the background noise in >50% of samples were discarded, (*iii*) the probe with the most consistent pattern of regional variation across donors was selected when more than one probe indexed the expression of the same gene, (*iv*) samples were assigned to a brain region if the coordinates were within 2 mm of the region boundary, and (*v*) gene expression was normalised across tissue samples.

We used a parcellation resulting from subdividing the Desikan–Killiany anatomical atlas into 316 cortical parcels of approximately equal surface area (around 500 mm^2^; https://github.com/RafaelRomeroGarcia/subParcellation_symmetric).^35^ All analyses were restricted to the left hemisphere (LH) because it was exhaustively covered in all donors. This procedure resulted in a 159 × 15634 (number of LH regions by genes) expression matrix describing the complete molecular profile of the normative LH.

### Constructing the grade-related expression map

The differential expression values derived from comparing GBM and LGG bulk tissue from the TCGA dataset were used as weights, where high positive weights correspond with genes that are over-expressed in GBM compared to LGG, and vice-versa. These weights were multiplied by the normative expression values across the cortex (ABHA data) to obtain a grade-related expression map. Thus, combining tumour mRNA and the spatial expression of genes in normative individuals, we derived an expression map representing brain regions that tend to express genes that are overexpressed in GBM compared to LGG. This map is hypothesised to reflect transcriptomic vulnerability to GBM. See purple box in **Figure 1** for a summary.

### Principal components of brain-related and tumour-related genes expression

The high co-expression between genes and the spatial distance effect of gene expression (i.e., closer regions tend to have similar expression profiles) implies that the dimensionality of the ABHA data can be effectively reduced to a few components which explain the vast majority of expression variability. We performed a Principal Component Analysis (PCA) over the normative gene expression matrix (159 regions x 15,634 genes) derived from the ABHA to obtain the most relevant regional expression patterns of brain-related genes. As in previous studies, we exploited the first two components (PCA1 and PCA2), resulting in a 159 regions by 2 PCA matrix revealing how each of the two gradients weight a given region.^36^ This linear dimensionality reduction technique allows visual and analytical exploration of the gene expression profiles.

Additionally, we identified a set of 15 genes derived from 25 loci that have been associated with tumour grading in a GWAS study^11^: IDH1, ATRX, TERT, MGMT, EGFR, PDGFRA1, TP53, NF1, MDM2, CDKN2A, CDKN2B, PTEN, PIK3CA, MYCN, CIC, FUBP1, NOTCH1and PI3K. The same PCA procedure was performed over this set of genes (i.e., using a 159 regions x 15 genes expression matrix) to derive the most relevant regional expression patterns of tumour-related genes. See green box in **Figure 1** for a summary.

### Brain network attributes

The 35,830 individuals that were available and pre-processed by the UK Biobank at the time of these analyses were used to derive robust brain connectivity markers in normative participants. Pre-processing included motion correction, intensity normalisation, high pass temporal filtering, EPI unwarping and artefacts removal by ICA+FIX processing, and are described elsewhere.^37^ Parcellations were transformed from structural T1-weighted image space to functional (EPI) MRI space using linear transformations. Regional time-series were computed as the average time-series of all voxels in the grey matter of each region. Functional connectivity between regions was calculated as the Pearson correlation between regional time-series.

Nodal strength was calculated for each individual as the average functional connectivity, representing regions that are more strongly functionally synchronized with other regions (hubs). As nodal strength does not take into account the community structure of the brain and is influenced by community size,^38^ we additionally computed participation coefficients (PC) as, 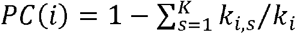 where *K* is the number of communities, *k*_*i*_ is the degree of the node *i*, and *k*_*i,s*_ is the intra-community degree of node *i* (representing the inter-modular connections of each node). The PC captures the nodes that facilitate communication between communities that make up the brain network. Finally, canonical (group-level) nodal strength and PC indices used in further analyses were calculated as the average regional values across all individuals. See red box in **Figure 1** for a summary.

### Statistical analyses

Associations were calculated using Pearson correlation when the Kolmogorov–Smirnov tests retained the hypothesis that both the independent and dependent variable arises from standard normal distributions. Spearman’s Rho correlation was used otherwise.

Both traditional parametric and non-parametric methods for relating brain maps ignore the inherent spatial auto-correlation of brain features (i.e., the data independence assumption is violated). To avoid inflated estimations of significance values, correspondence among regional maps (i.e., gene expression, glioma frequency and network metrics) was statistically tested by generating 10,000 random rotations (i.e., spins) of the cortical parcellation to estimate the distribution of r-values (or rho values for Spearman’s correlations) under the null hypothesis. This process provided a reference (null-)distribution for significance testing (*P*_*spin*_) of brain feature associations across regions while controlling for spatial contiguity of the cortical surface.^39^ Given that brain lobes have characteristic gene expression patterns, points on scatterplots (representing brain regions) were colour coded according to lobes: Frontal (green), Parietal (purple), Temporal (yellow), Occipital (light blue) and Insula (blue).

Two separate hypotheses were tested when exploring the associations between regional patterns of brain- and tumour-related gene expressions with functional brain network measures and grade-related expression maps: (i) that the associations between both brain-related and tumour-related gene expressions maps with network and grade-related markers across brain regions would be significantly different from zero (*P*_*spin*_, computed as described above); and, (ii) that the associations between tumour-related gene expression and each marker would be stronger (i.e., explained more variance) than between brain-related genes and each marker (*P*_*perm*_). As tumour-related genes were constituted by 15 genes with an associated co-expression value, each of the 10,000 permutations was calculated by randomly selecting 15 genes with similar co-expression (between 95% and 105% of the real co-expression value) and by correlating them with the regional marker. *P*_*perm*_ was then calculated as the proportion of randomly permuted cases that explained more variance than the observed set of tumour-related genes. *P*_*perm*_<0.05 was the threshold for significance.

## RESULTS

### Regional distributions of GBM and LGG are overlapping, but distinct

Demographic and clinical information regarding the 242 patients with glioma with matched MRI scans from TCIA is included in **Table 1**. Group-level GBM and LGG occurrence was obtained by concatenating glioma masks across patients (**Figure 1, yellow box**).

**Table 1.** Demographic and clinical variables for 242 patients with glioma from The Cancer Imaging Archive. Abbreviations: M=male; F=female; NA=not available; GBM=glioblastoma; LGG=low-grade glioma; IDH=isocitrate dehydrogenase; SD=standard deviation.

|  |  |
| --- | --- |
| Age (years) | 52.9 (15.2) |
| Gender (M/F/NA) | 133/107/2 |
| Clinical variables: |  |
| Grade (GBM/LGG) | 135/107 |
| Molecular subtype (IDH-wt/IDH-mut-1p19q-codel/IDH-mut-1p19q-noncodel/NA) | 124/27/61/30 |

GBM and LGG occurrences were heterogeneously distributed across the cortex with GBM predominantly located in insular and temporal cortices and LGG preferentially presenting in frontal and insular cortices (**Figure 2A**). Consequently, despite the spatial distribution of high and low grade tumours being significantly correlated (rho=0.54, *P*_*spin*_<10^−4^), a substantial part of the variance remains unexplained (**Figure 2B**). Maps were subtracted to construct the grade-related frequency map, a regional map reflecting the preferential occurrence of GBM compared with LGG. This map revealed a gradient of tumour grade occurrence across the cortex, with frontal and insular cortices having greater LGG occurrence and temporal and occipital cortices having greater GBM occurrence.

**Figure 2:**
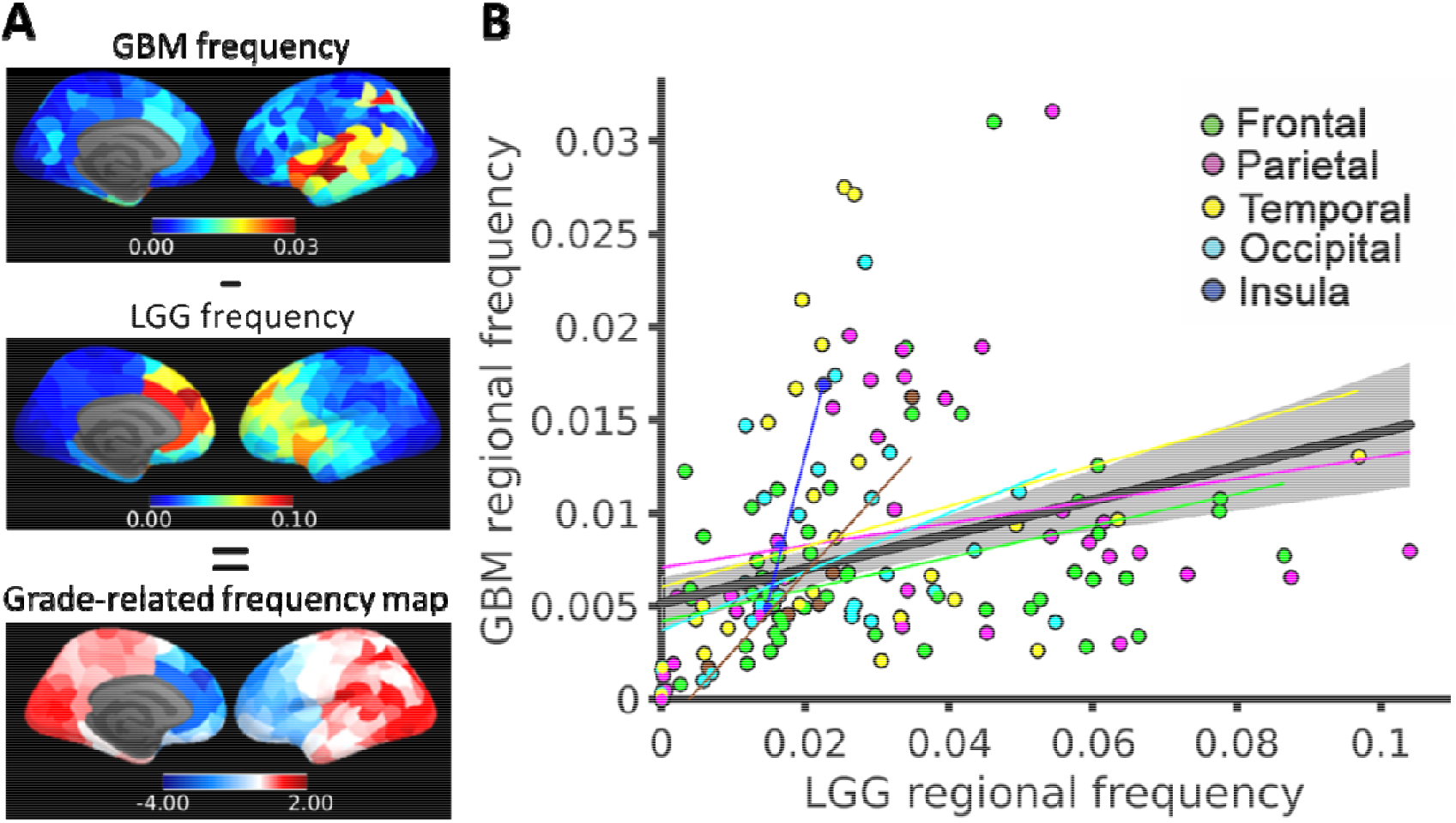
GBM and LGG regional distributions. **A)** Grade-related frequency map created from subtracting and z-scoring the GBM and LGG frequencies (i.e., occurrences) across brain regions. **B)** Association between LGG and GBM frequencies.

### Genes differentially expressed in GBM and LGG tissues show a characteristic expression pattern across brain regions

We initially determined how each gene of the genome was differentially expressed in GBM compared with LGG bulk tissue using the TCGA dataset (**Figure 1 purple box, Supplemental Data 1**). The top 1000 genes showing the highest differentiation between these two grades of glioma were significantly (P<10^−8^) enriched for neuronal elements, neurotransmitter activity and synaptic transmission by reference to the Gene Ontology resource (http://geneontology.org) (**Table 2, 3 and Supplemental Data 2**).

**Table 2.** Enrichment for molecular function of the top 1000 genes with greatest differential expression in GBM and LGG glioma tissues.

| Name | P-value | Adjusted p-value | Odds Ratio |
| --- | --- | --- | --- |
| voltage-gated cation channel activity (GO:0022843) | 3.03E-10 | 1.88E-07 | 6.02 |
| voltage-gated potassium channel activity (GO:0005249) | 1.59E-08 | 0.00000493 | 5.82 |
| ligand-gated cation channel activity (GO:0099094) | 3.43E-07 | 0.00007108 | 5.2 |
| GABA receptor activity (GO:0016917) | 5.19E-07 | 0.00008064 | 13.26 |
| calcium-dependent phospholipid binding (GO:0005544) | 7.13E-07 | 0.00008855 | 6.75 |
| GABA-gated chloride ion channel activity (GO:0022851) | 1.007E-06 | 0.00009956 | 22.32 |
| transmitter-gated ion channel activity involved in regulation of postsynaptic membrane potential (GO:1904315) | 1.122E-06 | 0.00009956 | 9.59 |
| GABA-A receptor activity (GO:0004890) | 1.753E-06 | 0.000136 | 13.92 |
| platelet-derived growth factor binding (GO:0048407) | 5.726E-06 | 0.0003556 | 22.93 |
| calcium ion binding (GO:0005509) | 5.467E-06 | 0.0003556 | 2.38 |
| potassium channel activity (GO:0005267) | 1.029E-05 | 0.0005808 | 4.1 |
| ligand-gated anion channel activity (GO:0099095) | 1.497E-05 | 0.0007749 | 12.17 |
| delayed rectifier potassium channel activity (GO:0005251) | 2.448E-05 | 0.001169 | 7.18 |
| cation channel activity (GO:0005261) | 2.697E-05 | 0.001196 | 3.75 |
| adenylate cyclase inhibiting G protein-coupled glutamate receptor activity (GO:0001640) | 3.293E-05 | 0.001278 | 23.86 |

**Table 3.** Enrichment for cellular components of the top 1000 genes with the greatest differential expression in GBM and LGG glioma tissues.

| Name | P-value | Adjusted p-value | Odds Ratio |
| --- | --- | --- | --- |
| neuron projection (GO:0043005) | 3.27E-17 | 8.87E-15 | 3.33 |
| collagen-containing extracellular matrix (GO:0062023) | 2.65E-14 | 3.59E-12 | 3.57 |
| integral component of plasma membrane (GO:0005887) | 2.38E-13 | 2.15E-11 | 2.13 |
| exocytic vesicle membrane (GO:0099501) | 4.70E-07 | 0.00002121 | 7.95 |
| synaptic vesicle membrane (GO:0030672) | 4.70E-07 | 0.00002121 | 7.95 |
| potassium channel complex (GO:0034705) | 3.43E-07 | 0.00002121 | 5.2 |
| GABA-A receptor complex (GO:1902711) | 1.753E-06 | 0.00006785 | 13.92 |
| dendrite (GO:0030425) | 2.017E-06 | 0.00006832 | 2.7 |
| voltage-gated potassium channel complex (GO:0008076) | 2.616E-06 | 0.00007877 | 4.97 |
| axon (GO:0030424) | 1.133E-05 | 0.0003072 | 2.82 |
| postsynaptic density membrane (GO:0098839) | 0.0000334 | 0.0008228 | 10.3 |
| postsynaptic specialization membrane (GO:0099634) | 4.794E-05 | 0.001083 | 9.56 |
| dense core granule (GO:0031045) | 7.994E-05 | 0.001667 | 11.46 |
| excitatory synapse (GO:0060076) | 0.0001663 | 0.003218 | 7.43 |
| cytoskeleton of presynaptic active zone (GO:0048788) | 0.0001926 | 0.003479 | 25.43 |

The tumour grade-related expression values were multiplied by the canonical gene expression values from the brain tissue of *post mortem* control donors in the AHBA to derive a weighted average of genes differentially expressed in GBM and LGG across regions (**Figure 3A**). The resulting grade-related expression map showed that genes over-expressed in GBM tissue are preferentially expressed in occipital and parietal cortices, while genes overexpressed in LGG tissue are expressed in anterior cingulate, motor, parahippocampal and entorhinal regions. This pattern was aligned with the two most important gradients of regional expression of 14,899 brain-related genes in healthy individuals as captured by PCA 1 (32% explained variance; rho = -0.40; *P*_*spin*_=0.006) and PCA 2 (25% explained variance; rho = 0.54; *P*_*spin*_=0.006) (**Figure 3B left panel and 3C**).

**Figure 3:**
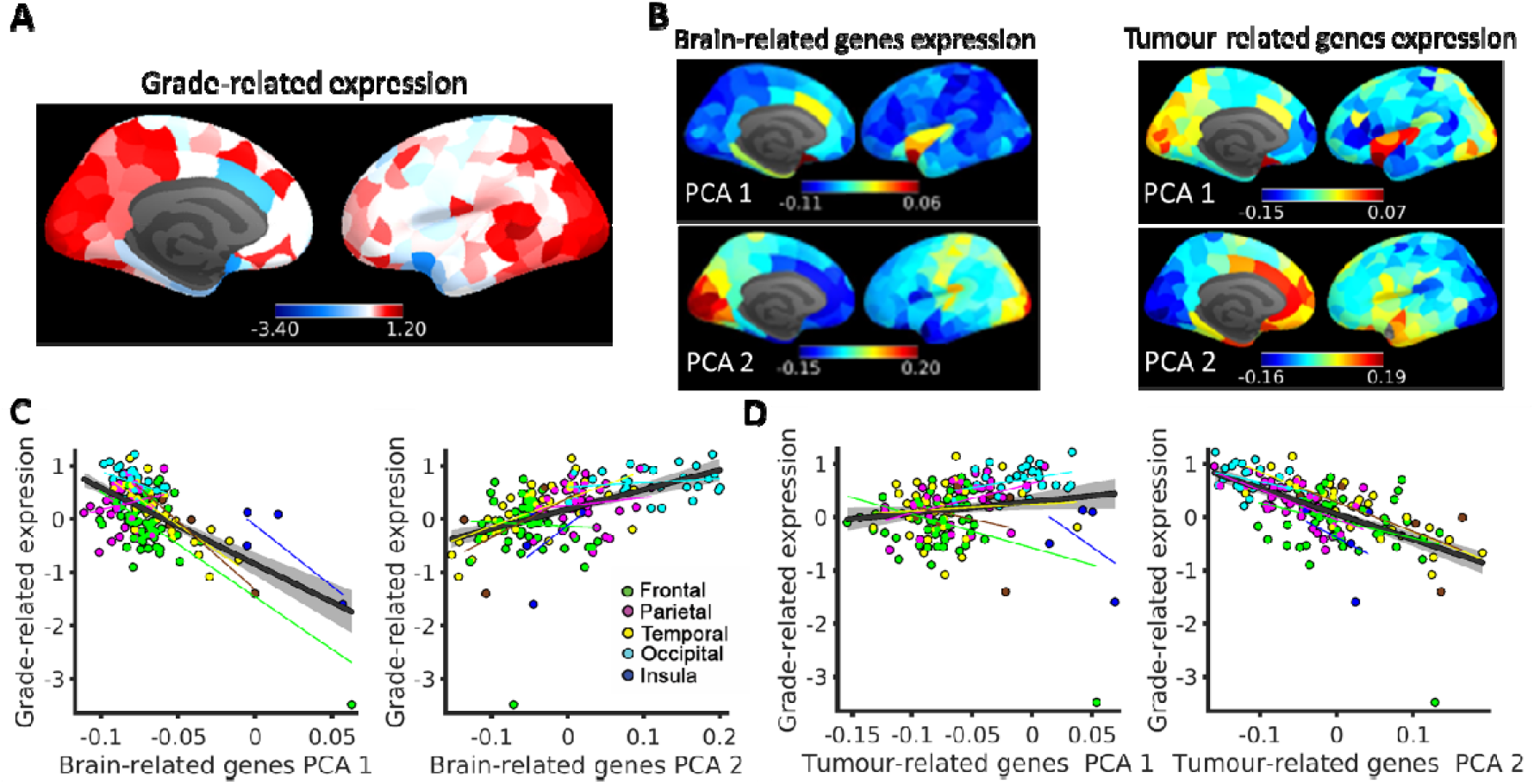
Regional pattern of genes differentially expressed in GBM and LGG tissues and tumour-related genes. **A)** Regional expression of brain-related genes in controls weighted by differential expression in GBM and LGG tissues (z-scored). **B)** Regional expression pattern derived from the ABHA of the first and second principal components of brain-related (left) and tumour-related (right) genes. **C)** Association between grade-related expression (shown in A) and the first and second principal components of brain-related genes from the ABHA. **D)** Association between grade-related expression and principal components of tumour-related gene expression.

Most genes that, according to a previous GWAS study^11^, present frequent molecular alterations on the five WHO 2016 subtypes of adult diffuse glioma (i.e. tumour-related genes) were significantly differentially expressed in GBM and LGG tissues (**Figure 1 green box**,13 out 15 were significant *P*_*fdr*_<0.01; **Supplemental Table S1**). When deriving the gene expression profiles of these genes from the ABHA dataset (**Figure 1 blue box**), we found two principal gradients of expression in healthy individuals: PCA 1 explaining 39% of the variance, PCA 2 explaining 16% of the variance; **Figure 3B right panel**. These principal components were also aligned with the grade-related expression map (PCA 1 - rho = 0.40; *P*_*spin*_=0.01; PCA 2 - rho = -0.65; *P*_*spin*_=0.0001; **Figure 3D**). Expression values were also computed comparing IDH1 wildtype and IDH1 mutated tumours, showing a similar differential expression profile (**Figure S1A**; R^2^=0.66, *P* ≃ 0) and spatial profile than when comparing GBM and LGG (**Figure S1B**; R^2^=0.87, *P*_*spin*_<10^−4^).

The tumour-related genes expression map significantly explained more variance of the grade-related gene expression map than randomly selected sets of brain-related genes under a null hypothesis (10,000 permutations preserving the number of tumour-related genes and their coexpression, *P*_*perm*_=0.003). Despite both expression maps being constructed using different approaches (i.e., considering genes differentially expressed in tumour tissue and GWAS-derived genes^11^), they both captured a similar transcriptional gradient of tumour grade vulnerability.

### Tumour grade-related frequency is associated with brain network features

We compared the glioma frequency maps with regional brain network measurements derived from healthy individuals of the UK Biobank dataset: nodal strength (i.e., regions that are more strongly functionally synchronized) and participation coefficient (i.e., regions with more inter-modular connections) (**Figure 1 red box**). The GBM frequency map was correlated with participation coefficient (rho = 0.24; *P*_*spin*_=0.0026), but not nodal strength (rho = 0.14; *P*_*spin*_=0.09; **Figure 4A**). Conversely, the LGG frequency map was significantly correlated with nodal strength (rho = -0.42; *P*_*spin*_=0.0001), but not participation coefficient (rho = 0.03; *P*_*spin*_=0.71; **Figure 4B**). This evidence indicates that regions playing a central role in healthy brain network topology may be key sites of vulnerability for aggressive tumours.

**Figure 4:**
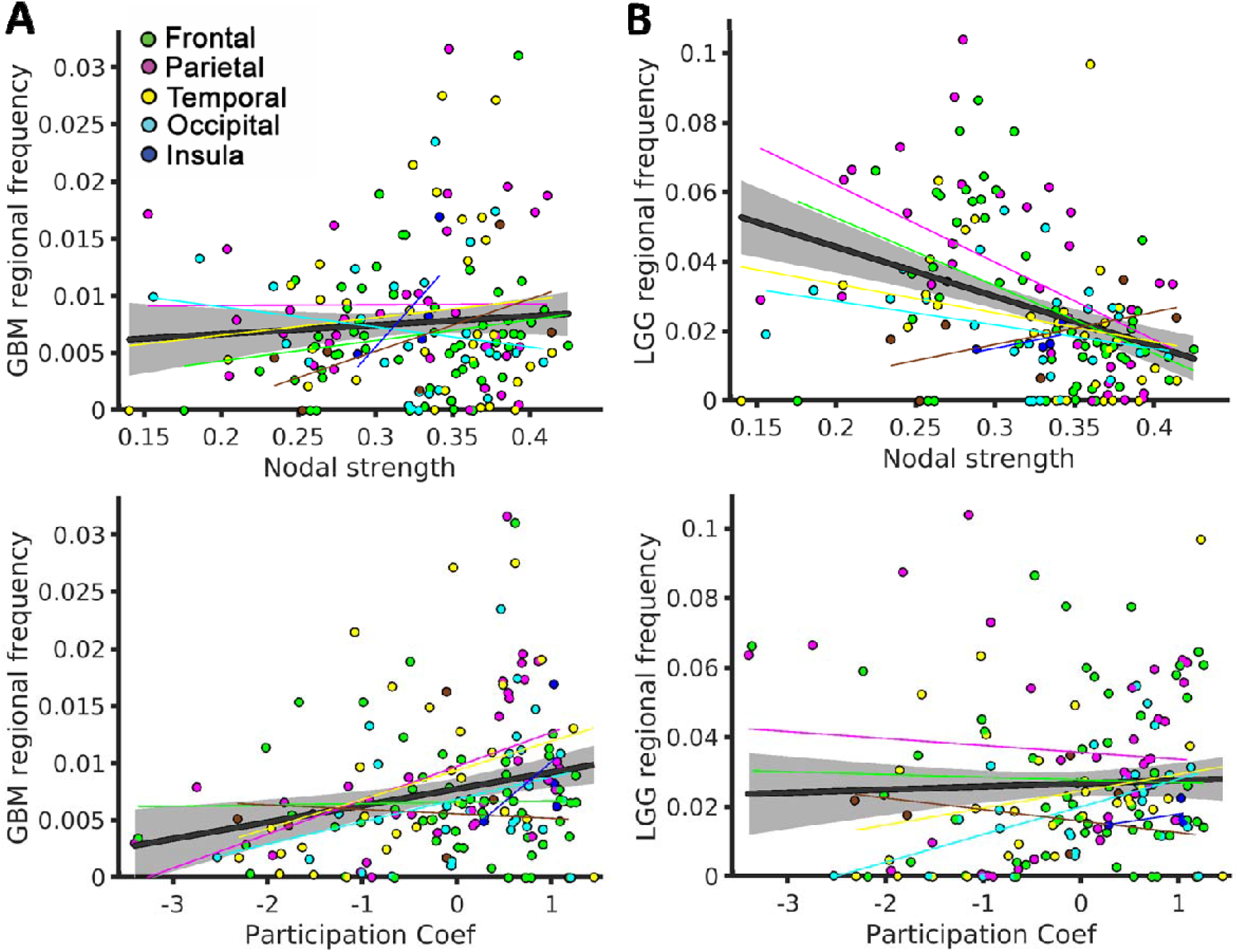
The regional distribution of GBMs and LGGs is associated with brain network features. **A)** Association between GBM regional frequency and nodal strength and participation coefficient. **B)** Association between LGG regional frequency and nodal strength and participation coefficient.

### The regional expression pattern of genes differentially expressed in GBM and LGG tissues is associated with grade-related frequency and brain network features

We next compared the regional patterns of the grade-related gene expression (**Figure 3**) with brain network features (**Figure 4**). Grade-related gene expression was not associated with glioma frequency (GBM and LGG occurrences combined; R^2^ =0.15; *P*_*spin*_=0.127), but was significantly associated with the grade-related frequency map (difference between GBM and LGG occurrences; rho = 0.51; *P*_*spin*_=0.038; **Figure 5A**). In other words, the grade-related gene expression pattern is related to the gradient of tumour grade occurrence across the cortex rather than with the total (pooled) tumour occurrence.

**Figure 5:**
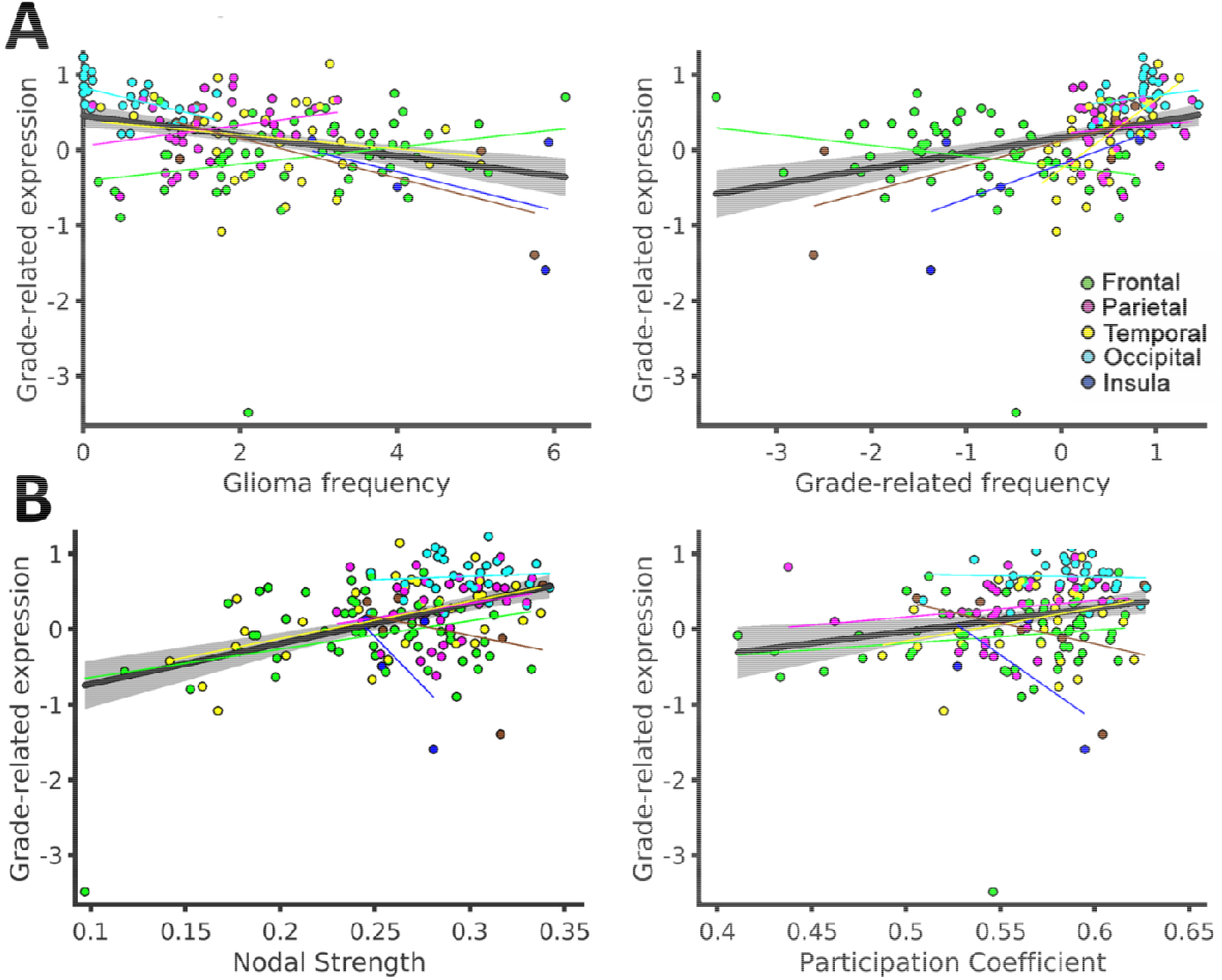
Genes differentially expressed in glioma tissue are associated with imaging markers. **A)** Grade-related gene expression association with glioma frequency (i.e., GBM and LGG occurrences combined) and grade-related frequency. **B)** Grade-related gene expression association with brain network nodal strength and participation coefficient.

When comparing the grade-related gene expression with brain network features we found that it was significantly associated with both nodal strength (R^2^ =0.11; *P*_*spin*_=0.006) and participation coefficient (R^2^ =0.07; *P*_*spin*_=0.016; **Figure 5B)**. These associations suggest that regional vulnerability to aggressive tumours might be partially mediated by network features and by the normative expression (in healthy brains) of genes over-expressed by these tumours.

### Expression of tumour-related genes is associated with grade-related frequency map and with nodal strength

The regional expression pattern of tumour-related genes was associated with grade-related frequency and brain network features. Specifically, grade-related frequency was significantly associated with tumour-related PCA 2 (rho = -0.64; *P*_*spin*_=0.003), but not PCA 1 (rho = 0.42; *P*_*spin*_=0.08) (**Figure 6A**). Similarly, PCA 2 but not PCA 1 was associated with both brain network features: nodal strength (**Figure 6B**) and participation coefficient (**Figure 6C**) (Nodal Strength – PCA 1, rho = 0.32; *P*_*spin*_=0.08; Nodal Strength – PCA 2, rho = -0.51; *P*_*spin*_=0.003; Participation Coefficient – PCA 1, rho = 0.17; *P*_*spin*_=0.12; Participation Coefficient – PCA 2, rho = -0.26; *P*_*spin*_=0.039).

**Figure 6:**
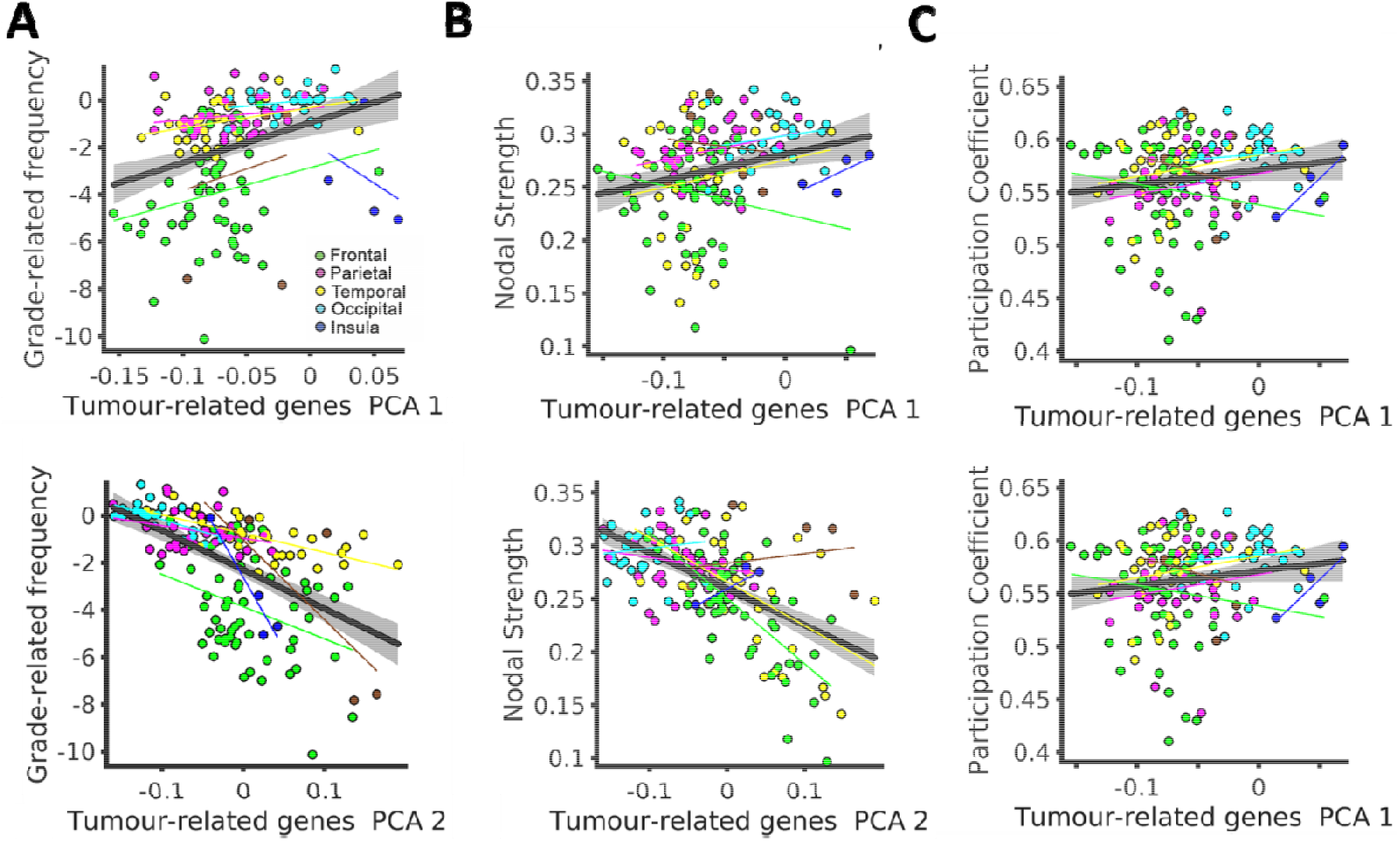
Associations between expression of grade-related frequency, tumour-related genes and brain network markers. **A)** Association between the two principal components of tumour-related gene expression and grade-related frequency. **B)** Association between the two principal components of tumour-related gene expression and network Nodal Strength. **C)** Association between the two principal components of tumour-related gene expression and network Participation Coefficient.

Associations were stronger for PCA 2 of tumour-related genes than for PCA 2 of brain-related genes (10,000 permutations; grade-related frequency *P*_*perm*_=0.006; Nodal Strength *P*_*perm*_=0.004; Participation Coefficient; *P*_*perm*_=0.004). This ordering of effect sizes in the relationships between tumour-related gene expression and network features, and grade-related frequency recapitulate our findings using the gene set derived by GWAS (**Figure 3**).

## DISCUSSION

Glioma occurrence across the cortex is not random, but is greater in frontal, temporal and insular lobes.^40,41^ From prior work that identified both genetics and brain function as key factors determining the spatial distribution of glioma,^19^ we hypothesized that they would similarly influence the differential pattern of occurrence related to tumour grade. Indeed, the transcriptomic and connectomic vulnerability factors in tumour emergence remain incompletely explored^42^.

We have shown that the pattern of differentially expressed genes in GBM and LGG tissues matches the corresponding frequency of their differential (i.e. grade-related) occurrence. Furthermore, the pattern of differentially expressed genes is also related to the topology and magnitude of the functional connectivity network, and to the expression of well-established tumour-related genes in normative controls. In combination, we found significant associations between functional connectivity, tumour-related gene expression and grade-related glioma frequency.

The interaction between glioma and neuronal elements has been known for over 80 years since HJ Scherer noted the predilection for gliomas to grow along and around normal neurons.^43^ Using MRI, we found that the GBM, but not the LGG frequency distribution was significantly associated with the participation coefficient of functional networks in a way that suggests that bridges inter-connecting constituent communities are more vulnerable to the appearance of malignant tumours. Interestingly functional network architecture, particularly regions with high connectivity, has been reproducibly linked with structural changes characteristic of psychiatric and neurological diseases.^44,45^

An extensive literature demonstrates that gliomas are electrically and synaptically integrated into neural circuits. Micro-scale experimental studies in animal models indicate that electrochemical communication occurs through bona fide AMPA receptor-dependent neuron-glioma synapses.^46^ At the macroscale circuit level, Numan et al. (2022) describes gliomas with increased malignancy preferentially occurring in regions characterized by higher brain activity in human controls.^47^ Functional brain networks have also been implicated as the substrate for structural lesions in a wide variety of psychiatry disorders, and appear to influence the location of primary tumours, albeit with a reduced effect size relative to genetic co-expression.^15^ Additionally, tumour functional integration within the global brain signal has been linked with cognitive recovery after glioma surgery.^48^ This study demonstrates that the principle of intimate entanglement between brain networks and patterns of neuropathological change is also relevant to tumour emergence and development.

Understanding the interplay between the molecular alterations that occur in gliomas and the transcriptomic signature of the normative brain identifies vulnerability factors that help explain the origins and progression of tumours. Here, we found that the differential expression profiles between GBM and LGG tissues were associated with the expression of brain-related genes and tumour-related genes (previously identified in the literature) in normative controls. The association with brain-related genes is likely mediated by cell type as the main contributing factor of the regional expression profile.^49^ Accordingly, Tan et al. (2013) reported that the first PCA component of the ABHA dataset is composed of two anti-correlated patterns enriched in oligodendrocyte and neuronal markers, respectively^50^, a pattern that has been also described in the mouse brain.^51^ The potential role of neuronal components in the distribution of gliomas is additionally supported by the enrichment of neuronal elements, neurotransmitter activity and synaptic transmission that were found in the gene ontology analysis of the GBM and LGG tissues. Variance of the grade-related gene expression map was significantly better explained by PCA2 expression patterns for tumour-related than for brain-related genes, indicating that cell types are only one contributing factor.

Transcriptomic risk factors of glioma occurrence are poorly understood due to limited data on healthy brain tissue. Conversely, the genetic vulnerability of glioma has been widely studied. The contribution of environmental factors is small (except for moderate to high doses of ionising radiation) compared with genetic risk.^52^ For example, first-degree relatives of glioma patients have a twofold increased risk of developing primary brain tumours compared with first-degree relatives of unaffected individuals.^53,54^ Genetic vulnerability does not only influence the risk of glioma appearance, but also the susceptibility to GBM and non-GBM tumours, reflecting their different aetiologies.^10^ How a molecular signature impacts prognosis is still a matter of debate. It has been proposed that specific molecular profiles allow more time for neuroplastic reorganisation, reducing the “lesion momentum” and improving not only survival rates, but also having a protective effect on neurocognitive functioning.^55^

Using similar approaches that exploit the ABHA dataset, a number of studies have shown regional expression patterns in normative controls that can be linked to structural and functional brain alterations of disease-related genes in psychiatric^24,25,56^ and neurological^57,58^ conditions. Here we extended, for the first time, this principle of transcriptomic vulnerability to neurooncology by showing that grade-related occurrence was associated with the expression in controls of tumour-related genes and genes that were differentially expressed in GBM and LGG.

Despite the discovery by GWAS of 25 susceptibility variants associated with GBM and non-GBM tumours, the expression profile across the normative cortex of the associated genes remains to be explored. Characterizing the transcriptomic signature is particularly important for understanding causal genetic associations in this context because the majority of the 25 loci reside in non-coding regions.^10^ Complementarily, a recent transcriptomic-wide association study identified 31 genes differentially expressed in GBM and non-GBM tumours,^59^ highlighting the important role that expression regulation has on tumour grade. For this reason, here we augmented the analysis of tumour-related genes (identified by the 25 loci) with a gene expression weighted average based on the differential expression between GBM and LGG bulk tissues. We found not only a significant association between both regional expression patterns, but also with connectivity features.

What emerges from these investigations is that a correspondence between tumour and brain genetic expression is an important factor in the instantiation of tumour cells at particular locations in the cerebral cortex. This goes some way to explaining the heterogeneous distribution of GBM and LGG which has a clear anterior-posterior gradient in differential frequency of occurrence (Figure 2). A large component of variance of the grade-related gene expression was significantly related to the grade-related frequency map (Figure 5), which was supported by the significant association of the regional expression pattern of tumour-related genes with grade-related frequency (Figure 6). Thus, as an overall motivating framework the seed-and-soil hypothesis, first suggested for brain metastases, could also be invoked for primary tumours, with the genetic signature of precursor cells (seed), and later the tumour itself, preferentially locating in brain regions with an appropriate gene expression (soil).

The success of this framework depends on the accommodation of other well-replicated factors that lie beyond the scope of this article. Age is a key factor, with GBM typically occurring in older adults relative to LGG, and a higher incidence in males relative to females.^60^ Although genes are crucial to understanding cancer and its treatment, our comprehension of genetic expression in brain remains incomplete and at an early stage. Nevertheless, it appears that there is variation of gene expression between the sexes^61^ and across the lifespan in a sex-dependent manner.^62^ As further, more detailed genetic data become available it should be possible to test if these broad differences are sufficient to explain the observed patterns as part of a seed-and-soil approach.

The reported frequency of LGGs undergoing malignant transformation ranges from 25% to 72%.^63,64^ Based on the analyses here, and if the seed-and-soil hypothesis is adopted then we can hypothesise that the probability of transformation is dependent on the degree of grade-related gene co-expression between tumour and brain. Circumstantial evidence for this comes from the identification of molecular classification of the tumour as well as male sex as risk factors for transformation.^63^ Our hypothesis would suggest that transforming tumours will be heterogeneously distributed across the brain, although to our knowledge this has not yet been measured.

Overall, given the myriad of studies that have linked brain conditions with normative expression, it is likely that the complex genetic architecture of brain diseases points toward a combined effect of genetic variants and transcriptomic factors that underlie regional brain vulnerability.^36^

### Limitations

The main limitation of combining datasets from normative individuals (UK Biobank and ABHA) is that templates do not contain information about brains of glioma patients (TCGA dataset). As a result, neurotypical functional networks and regional gene expression maps are averages of a normative sample and are not sensitive to intrinsic interindividual differences, omitting important tumour-specific idiosyncrasies.^42^

To maintain statistical power, tumours were not subclassified according to their molecular signature. The brain tissue samples used for RNA sequencing in the AHBA were not homogeneously distributed across the cortex, so estimates of regional expression are based on different numbers of experimental measurements at each of the 159 regions. However, equally sized regions were used to minimise the heterogeneity of sample distribution. Additionally, samples could not be matched across datasets for changes across the life span of gene expression^65^ profiles and functional connectivity^66^.

### Conclusions

Regional vulnerability to glioma frequency of occurrence is associated with normative brain expression patterns of tumour-related genes and grade-related differentially expressed genes. Moreover, this tumour grade-related regional vulnerability was associated with features of the functional connectivity network suggesting an interaction between tumour molecular, histological and functional brain architecture. Despite the limitations of establishing associations between multimodal markers derived from individuals with different demographic and clinical profiles, our study demonstrates the potential of combining transcriptomic data with MRI for a richer understanding of the neuropathological processes disrupting brain functioning in patients that can adversely affect quality of life.

## Data Availability

All data used is publicly available. Anonymized lesion data for GBM and LGG, respectively, are available at: https://www.cancerimagingarchive.net/access-data/. Clinical and genomic data from TCGA can be downloaded in R by following the workflow described in https://www.bioconductor.org/packages/release/workflows/vignettes/TCGAWorkflow/inst/doc/TCGAWorkflow.html. UK BioBank neuroimaging data are available at: https://www.fmrib.ox.ac.uk/ukbiobank/.

## ACKNOWLEDGEMENTS

We thank TCIA and TCGA for access to the neuroimaging and genomic data used in this study, as well as the patients who participated in those projects. This research also relied on imaging data from UK Biobank and The Allen Brain Institute.

## REFERENCES

1. Kumthekar P, Raizer J, Singh S. Low-grade glioma. Cancer Treat Res. Published online 2015. doi:10.1007/978-3-319-12048-5_5

2. Chang K, Zhang B, Guo X, et al. Multimodal imaging patterns predict survival in recurrent glioblastoma patients treated with bevacizumab. Neuro Oncol. Published online 2016. doi:10.1093/neuonc/now086

3. Walid M. Prognostic Factors for Long-Term Survival after Glioblastoma. Perm J. Published online 2008. doi:10.7812/tpp/08-027

4. Ohgaki H, Kleihues P. The definition of primary and secondary glioblastoma. Clin Cancer Res. Published online 2013. doi:10.1158/1078-0432.CCR-12-3002

5. Louis DN, Perry A, Reifenberger G, et al. The 2016 World Health Organization Classification of Tumors of the Central Nervous System: a summary. Acta Neuropathol. Published online 2016. doi:10.1007/s00401-016-1545-1

6. Ceccarelli M, Barthel FP, Malta TM, et al. Molecular Profiling Reveals Biologically Discrete Subsets and Pathways of Progression in Diffuse Glioma. Cell. 2016;164(3):550–563. doi:10.1016/j.cell.2015.12.028

7. Baldock AL, Ahn S, Rockne R, et al. Patient-specific metrics of invasiveness reveal significant prognostic benefit of resection in a predictable subset of gliomas. PLoS One. 2014;9(10). doi:10.1371/journal.pone.0099057

8. Tejada Neyra MA, Neuberger U, Reinhardt A, et al. Voxel-wise radiogenomic mapping of tumor location with key molecular alterations in patients with glioma. Neuro Oncol. Published online 2018. doi:10.1093/neuonc/noy134

9. Chen R, Nishimura MC, Kharbanda S, et al. Hominoid-specific enzyme GLUD2 promotes growth of IDH1R132Hglioma. Proc Natl Acad Sci U S A. Published online 2014. doi:10.1073/pnas.1409653111

10. Melin BS, Barnholtz-Sloan JS, Wrensch MR, et al. Genome-wide association study of glioma subtypes identifies specific differences in genetic susceptibility to glioblastoma and non-glioblastoma tumors. Nat Genet. Published online 2017. doi:10.1038/ng.3823

11. Molinaro AM, Taylor JW, Wiencke JK, Wrensch MR. Genetic and molecular epidemiology of adult diffuse glioma. Nat Rev Neurol. Published online 2019. doi:10.1038/s41582-019-0220-2

12. Ramakrishna R, Rostomily R. Seed, soil, and beyond: The basic biology of brain metastasis. Surg Neurol Int. Published online 2013. doi:10.4103/2152-7806.111303

13. Paget S. THE DISTRIBUTION OF SECONDARY GROWTHS IN CANCER OF THE BREAST. Lancet. Published online 1889. doi:10.1016/S0140-6736(00)49915-0

14. Sanai N, Alvarez-Buylla A, Berger MS. Neural Stem Cells and the Origin of Gliomas. N Engl J Med. Published online 2005. doi:10.1056/nejmra043666

15. Mandal AS, Romero-Garcia R, Seidlitz J, Hart MG, Alexander-Bloch AF, Suckling J. Lesion covariance networks reveal proposed origins and pathways of diffuse gliomas. Brain Commun. Published online 2021. doi:10.1093/braincomms/fcab289

16. Gillespie S, Monje M. An active role for neurons in glioma progression: Making sense of Scherer’s structures. Neuro Oncol. Published online 2018. doi:10.1093/neuonc/noy083

17. Monje M. Synaptic communication in brain cancer. Cancer Res. Published online 2020. doi:10.1158/0008-5472.CAN-20-0646

18. Campbell SL, Buckingham SC, Sontheimer H. Human glioma cells induce hyperexcitability in cortical networks. Epilepsia. Published online 2012. doi:10.1111/j.1528-1167.2012.03557.x

19. Mandal AS, Romero-garcia R, Hart MG, Suckling J. Genetic, cellular, and connectomic characterization of the brain regions commonly plagued by glioma. Brain. Published online 2020:3294-3307. doi:10.1093/brain/awaa277

20. Comprehensive, Integrative Genomic Analysis of Diffuse Lower-Grade Gliomas. N Engl J Med. Published online 2015. doi:10.1056/nejmoa1402121

21. Clark K, Vendt B, Smith K, et al. The cancer imaging archive (TCIA): Maintaining and operating a public information repository. J Digit Imaging. Published online 2013. doi:10.1007/s10278-013-9622-7

22. Hawrylycz MJ, Lein ES, Guillozet-Bongaarts AL, et al. An anatomically comprehensive atlas of the adult human brain transcriptome. Nature. 2012;489:391–39.

23. Romero-Garcia R, Seidlitz J, Whitaker KJ, et al. Schizotypy-Related Magnetization of Cortex in Healthy Adolescence Is Colocated With Expression of Schizophrenia-Related Genes. Biol Psychiatry. 2020;88(3):248–259. doi:10.1016/j.biopsych.2019.12.005

24. Romero-Garcia R, Hook RW, Tiego J, et al. Brain micro-architecture and disinhibition: a latent phenotyping study across 33 impulsive and compulsive behaviours. Neuropsychopharmacology. Published online January 1, 2020. doi:10.1038/s41386-020-00848-9

25. Romero-Garcia R, Warrier V, Bullmore ET, Baron-Cohen S, Bethlehem RAI. Synaptic and transcriptionally downregulated genes are associated with cortical thickness differences in autism. Mol Psychiatry. 2019;24(7):1053–1064. doi:10.1038/s41380-018-0023-7

26. Sudlow C, Gallacher J, Allen N, et al. UK Biobank: An Open Access Resource for Identifying the Causes of a Wide Range of Complex Diseases of Middle and Old Age. PLoS Med. Published online 2015. doi:10.1371/journal.pmed.1001779

27. McLendon R, Friedman A, Bigner D, et al. Comprehensive genomic characterization defines human glioblastoma genes and core pathways. Nature. Published online 2008. doi:10.1038/nature07385

28. Schmainda K, Prah M. The Cancer Imaging Archive. Published online 2018. doi:https://doi.org/10.7937/K9/TCIA.2018.15quzvnb

29. Silva TC, Colaprico A, Olsen C, et al. TCGA Workflow: Analyze cancer genomics and epigenomics data using Bioconductor packages. F1000Research. Published online 2016. doi:10.12688/f1000research.8923.2

30. Robinson MD, McCarthy DJ, Smyth GK. edgeR: A Bioconductor package for differential expression analysis of digital gene expression data. Bioinformatics. Published online 2009. doi:10.1093/bioinformatics/btp616

31. Bakas S, Akbari H, Sotiras A, et al. Advancing The Cancer Genome Atlas glioma MRI collections with expert segmentation labels and radiomic features. Sci Data. Published online 2017. doi:10.1038/sdata.2017.117

32. Chen EY, Tan CM, Kou Y, et al. Enrichr: interactive and collaborative HTML5 gene list enrichment analysis tool. BMC Bioinformatics. 2013;14(1):128. doi:10.1186/1471-2105-14-128

33. Romero-Garcia R, Whitaker KJ, Váša F, et al. Structural covariance networks are coupled to expression of genes enriched in supragranular layers of the human cortex. Neuroimage. 2018;171:256–267. doi:10.1016/j.neuroimage.2017.12.060

34. Markello RD, Arnatkeviciute A, Poline JB, Fulcher BD, Fornito A, Misic B. Standardizing workflows in imaging transcriptomics with the abagen toolbox. Elife. 2021;10. doi:10.7554/eLife.72129

35. Romero-Garcia R, Atienza M, Clemmensen LH, Cantero JL. Effects of network resolution on topological properties of human neocortex. Neuroimage. 2012;59(4):3522–3532. doi:10.1016/j.neuroimage.2011.10.086

36. Keo A, Mahfouz A, Ingrassia AMT, et al. Transcriptomic signatures of brain regional vulnerability to Parkinson’s disease. Commun Biol. Published online 2020. doi:10.1038/s42003-020-0804-9

37. Alfaro-Almagro F, Jenkinson M, Bangerter NK, et al. Image processing and Quality Control for the first 10,000 brain imaging datasets from UK Biobank. Neuroimage. Published online 2018. doi:10.1016/j.neuroimage.2017.10.034

38. Power JD, Schlaggar BL, Lessov-Schlaggar CN, Petersen SE. Evidence for hubs in human functional brain networks. Neuron. Published online 2013. doi:10.1016/j.neuron.2013.07.035

39. Váša F, Seidlitz J, Romero-Garcia R, et al. Adolescent tuning of association cortex in human structural brain networks. Cereb Cortex. Published online 2018. doi:10.1093/cercor/bhx249

40. Larjavaara S, Mäntylä R, Salminen T, et al. Incidence of gliomas by anatomic location. Neuro Oncol. Published online 2007. doi:10.1215/15228517-2007-016

41. Duffau H, Capelle L. Preferential brain locations of low-grade gliomas. Cancer. Published online 2004. doi:10.1002/cncr.20297

42. Germann J, Zadeh G, Mansouri A, Kucharczyk W, Lozano AM, Boutet A. Untapped Neuroimaging Tools for Neuro-Oncology: Connectomics and Spatial Transcriptomics. Cancers. 2022;14(3). doi:10.3390/cancers14030464

43. Scherer HJ. Structural development in gliomas. Am J Cancer. Published online 1938. doi:10.1158/ajc.1938.333

44. Seeley WW, Crawford RK, Zhou J, Miller BL, Greicius MD. Neurodegenerative Diseases Target Large-Scale Human Brain Networks. Neuron. Published online 2009. doi:10.1016/j.neuron.2009.03.024

45. Crossley N a., Mechelli A, Scott J, et al. The hubs of the human connectome are generally implicated in the anatomy of brain disorders. Brain. 2014;137(8):2382–2395. doi:10.1093/brain/awu132

46. Venkatesh HS, Morishita W, Geraghty AC, et al. Electrical and synaptic integration of glioma into neural circuits. Nature. Published online 2019. doi:10.1038/s41586-019-1563-y

47. Numan T, Breedt LC, Maciel B de APC, et al. Regional healthy brain activity, glioma occurrence and symptomatology. medRxiv. Published online January 1, 2022:2022.01.14.22269290. doi:10.1101/2022.01.14.22269290

48. Romero-Garcia R, Hart MG, Bethlehem RAI, et al. Bold coupling between lesioned and healthy brain is associated with glioma patients’ recovery. Cancers (Basel). Published online 2021. doi:10.3390/cancers13195008

49. Cahoy JD, Emery B, Kaushal A, et al. A transcriptome database for astrocytes, neurons, and oligodendrocytes: A new resource for understanding brain development and function. J Neurosci. Published online 2008. doi:10.1523/JNEUROSCI.4178-07.2008

50. Tan PPC, French L, Pavlidis P. Neuron-enriched gene expression patterns are regionally anti-correlated with oligodendrocyte-enriched patterns in the adult mouse and human brain. Front Neurosci. Published online 2013. doi:10.3389/fnins.2013.00005

51. French L, Tan PPC, Pavlidis P. Large-scale analysis of gene expression and connectivity in the rodent brain: Insights through data integration. Front Neuroinform. Published online 2011. doi:10.3389/fninf.2011.00012

52. Kinnersley B, Labussière M, Holroyd A, et al. Genome-wide association study identifies multiple susceptibility loci for glioma. Nat Commun. Published online 2015. doi:10.1038/ncomms9559

53. Hemminki K, Tretli S, Sundquist J, Johannesen TB, Granström C. Familial risks in nervous-system tumours: a histology-specific analysis from Sweden and Norway. Lancet Oncol. Published online 2009. doi:10.1016/S1470-2045(09)70076-2

54. Malmer B, Grönberg H, Bergenheim AT, Lenner P, Henriksson R. Familial aggregation of astrocytoma in Northern Sweden: An epidemiological cohort study. Int J Cancer. Published online 1999. doi:10.1002/(SICI)1097-0215(19990505)81:3<366::AID-IJC9>3.0.CO;2-0

55. Wefel JS, Noll KR, Scheurer ME. Neurocognitive functioning and genetic variation in patients with primary brain tumours. Lancet Oncol. 2016;17(3):e97–e108. doi:10.1016/S1470-2045(15)00380-0

56. Romero-Garcia R, Seidlitz J, Whitaker KJ, et al. Schizotypy-Related Magnetization of Cortex in Healthy Adolescence Is Colocated With Expression of Schizophrenia-Related Genes. Biol Psychiatry. Published online 2020. doi:10.1016/j.biopsych.2019.12.005

57. Seidlitz J, Nadig A, Liu S, et al. Transcriptomic and cellular decoding of regional brain vulnerability to neurogenetic disorders. Nat Commun. 2020;11(1). doi:10.1038/s41467-020-17051-5

58. Kang S, Kim HR, Seong JK. Associating cognitive reserve and brain-wide gene expression regarding Alzheimer’s disease. Alzheimer’s Dement. 2021;17(S4):e049732. doi:https://doi.org/10.1002/alz.049732

59. Atkins I, Kinnersley B, Ostrom QT, et al. Transcriptome-wide association study identifies new candidate susceptibility genes for glioma. Cancer Res. Published online 2019. doi:10.1158/0008-5472.CAN-18-2888

60. Chakrabarti I, Cockburn M, Cozen W, Wang YP, Preston-Martin S. A population-based description of glioblastoma multiforme in Los Angeles County, 1974-1999. Cancer. 2005;104(12):2798–2806. doi:10.1002/CNCR.21539

61. Gegenhuber B, Tollkuhn J. Signatures of sex: Sex differences in gene expression in the vertebrate brain. Wiley Interdiscip Rev Dev Biol. 2020;9(1). doi:10.1002/WDEV.348

62. Berchtold NC, Cribbs DH, Coleman PD, et al. Gene expression changes in the course of normal brain aging are sexually dimorphic. Proc Natl Acad Sci U S A. 2008;105(40):15605–15610. doi:10.1073/PNAS.0806883105/SUPPL_FILE/ST9.XLS

63. Van Den Bent MJ, Afra D, De Witte O, et al. Long-term efficacy of early versus delayed radiotherapy for low-grade astrocytoma and oligodendroglioma in adults: the EORTC 22845 randomised trial. Lancet (London, England). 2005;366(9490):985–990. doi:10.1016/S0140-6736(05)67070-5

64. Chaichana KL, McGirt MJ, Laterra J, Olivi A, Quiñones-Hinojosa A. Recurrence and malignant degeneration after resection of adult hemispheric low-grade gliomas. J Neurosurg. 2010;112(1):10–17. doi:10.3171/2008.10.JNS08608

65. Qiu A, Zhang H, Kennedy BK, Lee A. Spatio-temporal correlates of gene expression and cortical morphology across lifespan and aging. Neuroimage. Published online 2021. doi:10.1016/j.neuroimage.2020.117426

66. Váša F, Romero-Garcia R, Kitzbichler MG, et al. Conservative and disruptive modes of adolescent change in human brain functional connectivity. Proc Natl Acad Sci U S A. Published online 2020. doi:10.1073/pnas.1906144117

