## Supplemental Information for "Transcriptomic and connectomic correlates of differential spatial patterning among glioblastomas and low-grade gliomas"

**SUPPLEMENTAL MATERIAL**

Rafael Romero-Garcia^1,2^, Ayan S. Mandal^2,3^, Richard AI Bethlehem^2^, Benedicto Crespo-Facorro^4^, Michael G Hart^5^, John Suckling^2,6,7^

1. Instituto de Biomedicina de Sevilla (IBiS) HUVR/CSIC/Universidad de Sevilla/ CIBERSAM, ISCIII, Dpto. de Fisiología Médica y Biofísica
2. Department of Psychiatry, University of Cambridge
3. Perelman School of Medicine, University of Pennsylvania
4. Hospital Universitario Virgen del Rocio, Department of Psychiatry, Instituto de Investigación Sanitaria de Sevilla, (IBiS), CIBERSAM, Sevilla, Spain
5. Neurosciences Research Centre, Institute of Molecular and Clinical Sciences, St George’s, University of London
6. Behavioural and Clinical Neuroscience Institute, University of Cambridge
7. Cambridge and Peterborough NHS Foundation Trust.

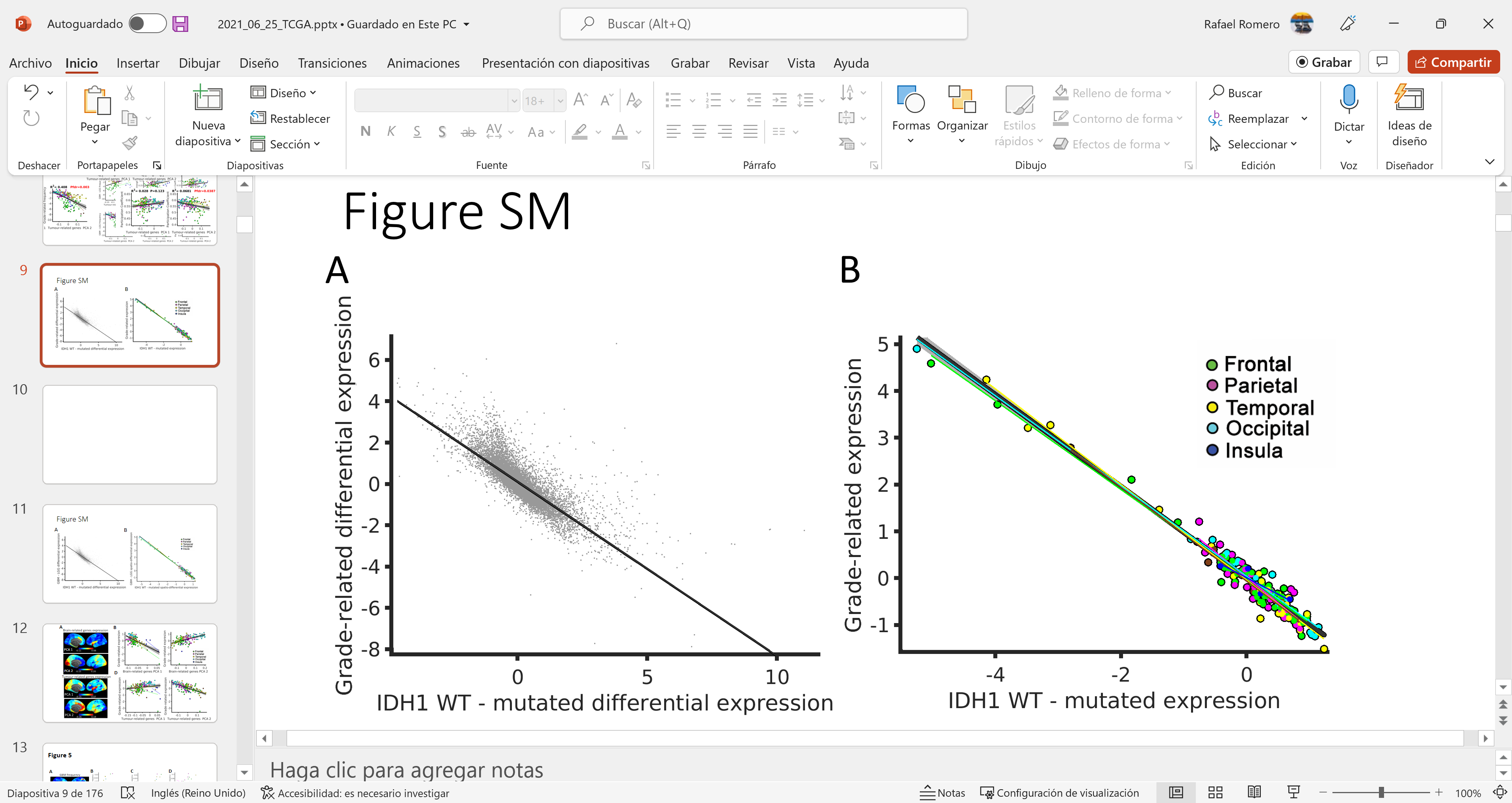

**Figure S1**. Association between expression values comparing IDH Wild Type vs. IDH1 Mutated (IDH-related) and GBM vs. LGG (Grade-related). (**A**) Association between differential expression values in bulk tissue (each point representing a gene). **B**) Association between spatial expression profiles in normative brain tissue (each point representing a brain region).

| **Gene name** | **logCPM** | **P-value**  **(FDR corrected)** |
| --- | --- | --- |
| IDH1 | -0.96 | 2.04E-14 |
| ATRX | 0.56 | 0.00164 |
| TERT | - | - |
| MGMT | -0.95 | 1.72E-06 |
| EGFR | -0.62 | 0.08395 |
| PDGFRA1 | - | - |
| TP53 | -0.65 | 1.67E-05 |
| NF1 | 1.15 | 3.03E-25 |
| MDM2 | -3.29 | 5.05E-29 |
| CDKN2A | -2.31 | 1.33E-10 |
| CDKN2B | 0.32 | 3.58E-01 |
| PTEN | 0.32 | 4.24E-04 |
| PIK3CA | 0.59 | 1.25E-05 |
| MYCN | 0.69 | 1.39E-02 |
| CIC | 0.12 | 2.64E-01 |
| FUBP1 | -0.08 | 0.5621 |
| NOTCH1 | 0.70 | 0.00011 |
| PI3K | - | - |

**Table S1**. Differential expression between GBM and LGG bulk tissue of tumour-related genes. – indicates genes where expression values were not available. logCPM represents log counts per million with positive values indicating higher expression in GBM compared to LGG.
